# Long-term follow-up of subthalamic nucleus deep brain stimulation in patients with Parkinson’s disease: an analysis of survival and disability milestones

**DOI:** 10.1101/2023.06.03.23290925

**Authors:** Raquel Barbosa, Leonor Correia Guedes, Maria Begoña Cattoni, Patricia Pita Lobo, Ana Castro Caldas, Margherita Fabbri, Paulo Bastos, Anabela Valadas, Herculano Carvalho, Luisa Albuquerque, Sofia Reimão, A Gonçalves Ferreira, Joaquim J Ferreira, Mário Miguel Rosa, Miguel Coelho

## Abstract

**Background:** Data on the long-term survival and incidence of disability milestones after subthalamic nucleus deep brain stimulation (STN-DBS) in Parkinson’s disease (PD) is limited.

**Objectives:** To estimate mortality among PD patients 8 years after STN-DBS and to quantify the frequency /time-to-development of disability milestones (falls, freezing, hallucinations, dementia, and institutionalization).

**Methods:** A longitudinal retrospective study of patients submitted to STN-DBS between 2006-2012 was carried-out. For mortality, Cox proportional hazards regression analysis was performed. For disease milestones, competing risk analyses were performed and cumulative incidence functions calculated. Multivariable regression analysis in the presence of competing risks was performed based on the Gray’s test of sub-distribution hazards of cumulative incidence functions.

**Results:** An overall mortality rate of 16% (mean 62.1 ± 21.3 months after surgery) was observed. Falls (73%) and freezing (47%) were both the earliest (40.4 ± 25.4 and 39.6 ± 28.4 months, respectively) and most frequently observed milestones. Dementia (34%) and hallucinations (32%) soon follow (56.2 ± 21.2 and mean 60.0 ± 20.7 months after surgery, respectively). Institutionalization occurred in 6% after 62.3 ± 22.0 months. Higher ADL scores in the OFF state and higher age at surgery were associated with falls, freezing, dementia and institutionalization.

**Conclusions:** Long-term mortality rate is low after STN-DBS, with STN-DBS patients developing the same disability milestones as non-DBS patients. Motor milestones occur first and at a higher frequency than cognitive ones. Milestones cluster together before death.

## Introduction

Deep brain stimulation (DBS) of the subthalamic nucleus (STN) is a well-recognized and effective treatment for patients with advanced Parkinson’s Disease (PD).^1^ Prior studies have shown that STN-DBS significantly improves levodopa-induced motor complications (MC), motor symptoms and quality-of-life (QoL) in the short-to-medium term (6 to 12 months and up to 5 years) ^2–8^ with a sustained benefit being observed in long-term follow-up studies (up to 15 years).^9–13^ However, data on the long-term survival and disability milestones is more limited.^14, 15^

Despite improvements in MC and appendicular signs upon STN-DBS, axial signs such as gait, freezing-of-gait (FOG), postural instability and dysarthria fail to experience comparable benefit in both medium- and long-term follow-ups,^10–12, 16^ probably reflecting disease progression and the involvement of non-dopaminergic pathways unresponsive to stimulation.^11–13, 16^ Indeed, studies on non-DBS patients have found that the emergence of these axial signs tend to cluster with the development of certain non-motor symptoms (e.g. psychosis, cognitive impairment, and autonomic dysfunction), and that such association is strongly correlated with disease progression.^17–21^ Accordingly, the development of dementia and hallucinations, occurrence of falls and nursing home placement represent important PD milestones that mirror disease progression, reflect disease severity and offer valuable insights into its prognosis. Additionally, gathering knowledge of the development of these disability milestones is important for assessing long-term safety of DBS. Of particular note, the concept of “late-stage disease” can be translated into this DBS population, whereby despite no longer disabled by tremor, rigidity or severe MC, patients still suffer from axial, dysautonomic, cognitive and behavioral symptoms non-responsive to stimulation (and dopaminergic) therapy that predominate the clinical picture.^17, 222^

Taking this into consideration, the aim of this study was to characterize the long-term survival of PD patients submitted to STN-DBS surgery and to identify the emergence of and the delay until the occurrence of disability milestones: falls, FOG, hallucinations, dementia, and institutionalization.

## Material & Methods

### *Design:* a longitudinal retrospective study

*Primary objective:* to assess mortality of PD patients submitted to bilateral STN-DBS after a follow-up of 8 years.

*Secondary objective:* to determine the frequency and the determinants of disability milestones (falls, FOG, hallucinations, dementia, and institutionalization) in the same population of patients.

*Patients:* All PD patients undergoing bilateral STN-DBS from 2006 to 2012 at Hospital Santa Maria were included in the study. Patients implanted in the *globus pallidus internus* were excluded (n=2). PD was diagnosed according to the UK Brain Bank criteria^23^ and the criteria for DBS were: i) presence of clinically significant levodopa-induced MC not optimally controlled with medication, ii)< 70 years-old, iii) a ≥ 33% reduction in the Unified Parkinson’s Disease Rating Scale (UPDRS)^24^ motor score after a supra-threshold levodopa dose, iv) no dementia, v) no major/refractory psychiatric illness, and vi) no postural instability or FOG in the best ON state. Patients who had the leads or the implanted pulse generator definitively removed during the follow-up were identified and censored at the time of DBS system removal.

*Study Outcomes*: *Primary outcome -* mortality during the 8 years after DBS. *Secondary outcomes -* occurrence of any of the following disability milestones: falls; FOG; hallucinations; dementia; and institutionalization.

All patient data was retrospectively retrieved and analysed from baseline and up to 8 years post-surgery. Data was extracted from every single hospital visit of patients. Patients were followed-up until the time of death or the end of the follow-up period (8 years), whichever occurred first.

### Assessment tools

Falls: defined as a fall report by the patient or caregivers on two consecutive visits.

FOG: defined by the presence of FOG observed by the clinician on two consecutive visits. Both falls and FOG were considered only after 6 months into the post-surgical period, as we considered 6 months the time required for treatment stabilization.

Dementia: defined according to the diagnostic criteria of the DSM-V.

Institutionalization: the date of placement in a nursing home was either the specific date recorded in files or the date of the visit when institutionalization was first mentioned.

Additional baseline pre-surgery data was collected: age and duration of disease at surgery, gender, scores of parts II (activities of daily living) and III (motor) of the UPDRS in both OFF and ON states, scores of part I (mental) and IV (motor complications) of UPDRS, Hoehn and Yahr (H&Y) stage^25^ and Schwab and England score in both ON and OFF state, percentage of response to a supra-threshold dose of levodopa. The percentage of patients with a significant gait impairment (score ≥2 on item 29 of the UPDRS-III) or postural instability (score ≥2 on item 30 of the UPDRS-III) in the OFF state were calculate. Clinical phenotype, i.e. postural instability/gait disorder (PIGD) and tremor dominant (TD), and levodopa-equivalent daily doses (LEDD) (mg/day) were calculated as previously described.^26, 27^ Data from formal neuropsychological evaluation including the MMSE score were collected.^28^ As our institution started using the MDS-UPDRS^29^ only from 2012 onwards, the MDS-UPDRS II and III scores were converted from the corresponding UPDRS ones using the available conversion formula,^30^ allowing for the standardization of the entire cohort.

## Data Analysis

Clinical and demographic characteristics were described as mean ± standard deviation or percentages, as appropriated. Two group comparisons were performed using Mann-Whitney U-test for continuous variables or Chi-square tests for categorical variables. Descriptive statistics and results from exploratory analysis for all variables of interest are in **Table 1**, **Supplementary Tables S1 and S2**. Multicollinearity was tested using Spearman coefficients between all possible pairs of variables (**Sup. Fig. S1**).

**Table 1.**
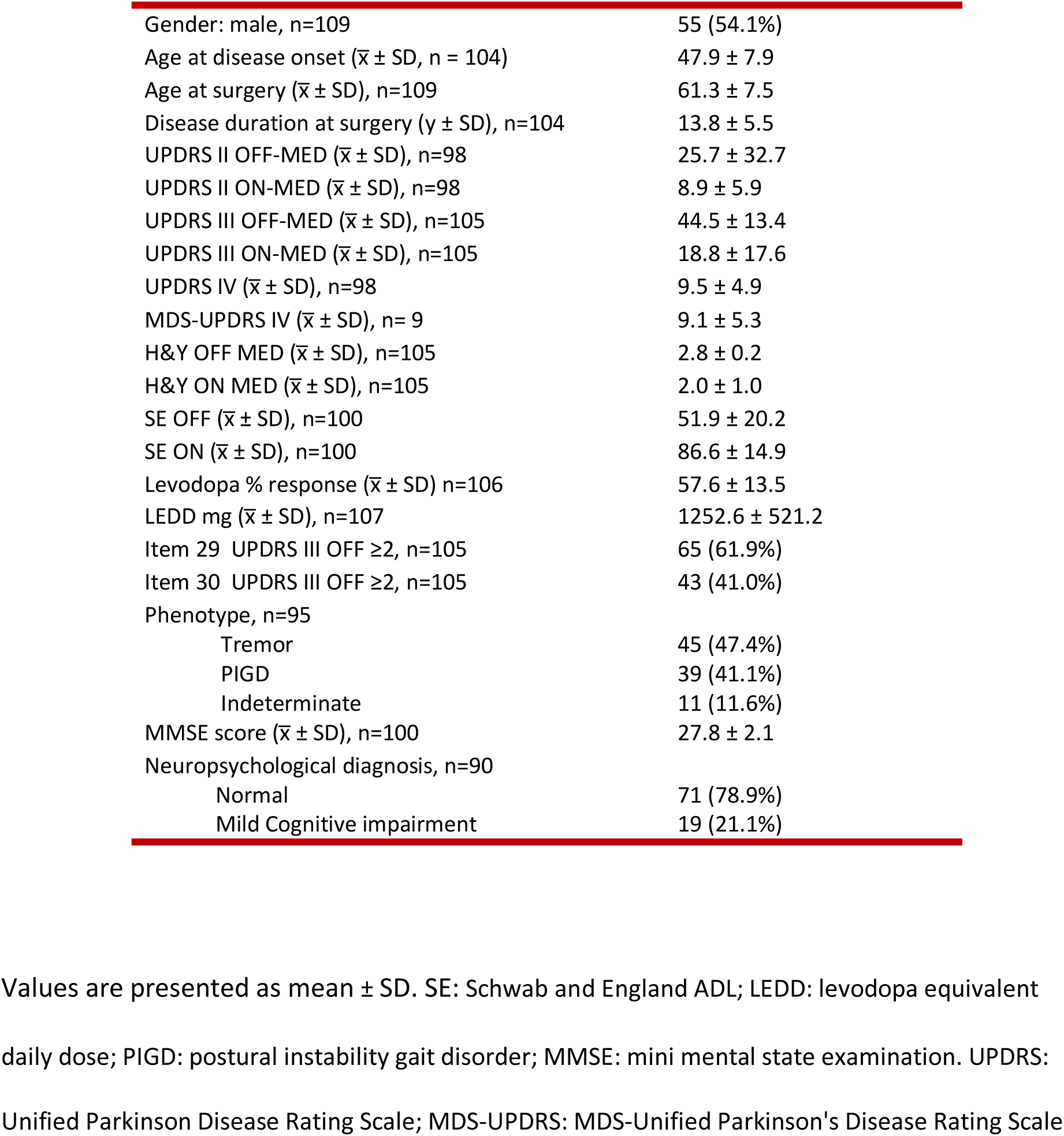
Demographic and clinic characteristics of study participants.

|  |  |
| --- | --- |
| Gender: male, n=109 | 55 (54.1%) |
| Age at disease onset ($\bar{x} \pm SD$, n = 104) | 47.9 ± 7.9 |
| Age at surgery ($\bar{x} \pm SD$), n=109 | 61.3 ± 7.5 |
| Disease duration at surgery ($y \pm SD$), n=104 | 13.8 ± 5.5 |
| UPDRS II OFF-MED ($\bar{x} \pm SD$), n=98 | 25.7 ± 32.7 |
| UPDRS II ON-MED ($\bar{x} \pm SD$), n=98 | 8.9 ± 5.9 |
| UPDRS III OFF-MED ($\bar{x} \pm SD$), n=105 | 44.5 ± 13.4 |
| UPDRS III ON-MED ($\bar{x} \pm SD$), n=105 | 18.8 ± 17.6 |
| UPDRS IV ($\bar{x} \pm SD$), n=98 | 9.5 ± 4.9 |
| MDS-UPDRS IV ($\bar{x} \pm SD$), n= 9 | 9.1 ± 5.3 |
| H&Y OFF MED ($\bar{x} \pm SD$), n=105 | 2.8 ± 0.2 |
| H&Y ON MED ($\bar{x} \pm SD$), n=105 | 2.0 ± 1.0 |
| SE OFF ($\bar{x} \pm SD$), n=100 | 51.9 ± 20.2 |
| SE ON ($\bar{x} \pm SD$), n=100 | 86.6 ± 14.9 |
| Levodopa % response ($\bar{x} \pm SD$) n=106 | 57.6 ± 13.5 |
| LEDD mg ($\bar{x} \pm SD$), n=107 | 1252.6 ± 521.2 |
| Item 29 UPDRS III OFF $\geq 2$, n=105 | 65 (61.9%) |
| Item 30 UPDRS III OFF $\geq 2$, n=105 | 43 (41.0%) |
| Phenotype, n=95 |  |
| Tremor | 45 (47.4%) |
| PIGD | 39 (41.1%) |
| Indeterminate | 11 (11.6%) |
| MMSE score ($\bar{x} \pm SD$), n=100 | 27.8 ± 2.1 |
| Neuropsychological diagnosis, n=90 |  |
| Normal | 71 (78.9%) |
| Mild Cognitive impairment | 19 (21.1%) |

Values are presented as mean ± SD. SE: Schwab and England ADL; LEDD: levodopa equivalent daily dose; PIGD: postural instability gait disorder; MMSE: mini mental state examination. UPDRS: Unified Parkinson Disease Rating Scale; MDS-UPDRS: MDS-Unified Parkinson's Disease Rating Scale

For the primary outcome, Cox proportional hazards regression analysis was performed to allow handling multiple quantitative and categorical variables simultaneously. The time-dependent survival was fit using each individual variable, and variables of interest were subsequently selected for statistical variable adjustment and multivariable regression. The log-rank (Mantel-Cox) test was used for comparison of the survival curves between groups and estimation of the respective hazard ratios (Mantel-Haenszel). For secondary outcomes, the primary outcome (i.e., death) hindered the occurrence of the secondary outcome in a time-dependent way. Thus, competing risk analysis were performed and cumulative incidence calculated for all secondary outcomes, as previously reported.^31^ Then, multivariable regression analysis in the presence of competing risks was performed using the semiparametric proportional hazards model (i.e. sub-distribution hazards of cumulative incidence functions) based on the Gray’s test, in order to assess the contributions of different variables (adjusted and non-adjusted) to the development of secondary outcomes.^32^ For all models, a backwards stepwise regression model was used, removing independent variables until all contributing variables had a p-value < 0.2. Analysis and graphical representations as cumulative incidence, survival or forest plots was performed using R 4.0.4 and the analysis pipeline can be found at [https://github.com/Le-bruit-de-nos-pas/PD_Milestones_STN-DBS]. Exact p-values, hazard ratios and 95% confident intervals are reported.

### Data availability statement

Anonymized data of this study will be available from the corresponding author on reasonable request from any qualified researcher, following the EU General Data Protection Regulation. analysis pipeline can be found at [https://github.com/Le-bruit-de-nos-pas/PD_Milestones_STN-DBS].

## Results

### Clinical and demographic baseline characteristics

109 patients underwent STN-DBS during the study period. Clinical and demographic data are presented in **Table 1**. During follow-up, 8 patients were censored: 7 due to system removal because of infection (at months 5, 35, 45, 57, 61, and 2x 95) and 1 was referred to another institution and lost to follow up (at month 72). 84 patients reached the end of the 8-year follow-up: mean age of 68.6 ± 7.6, mean disease duration of 21.38 ± 4.38 years, mean H&Y 2.8 ± 1.1, mean LEDD of 600.90 ± 370.14 mg (vs 1252.66 ± 521.17 mg at baseline; average reduction of 49.64 ± 29.52 %, *p* < 0.001). Stimulation parameters at the end of follow-up in **Table 2**.

**Table 2.** Stimulation parameters at the end of follow-up.

| Table 2 – Stimulation parameters at the end of follow-up |  |  |
| --- | --- | --- |
|  | Right STN (n=83) | Left STN (n=83) |
| Monopolar (%) | **81.9% (68)** | **85.5% (71)** |
| Voltage (mV ) | 3.91 ± 4.60 | 3.06 ± 0.47 |
| Pulse width (µs) | 63.1 ± 10.3 | 62.5 ± 9.67 |
| Frequency (Hz) | 123 ± 17.1 | 124 ± 17.1 |
| Bipolar (%) | **14.5% (12)** | **12.0% (10)** |
| Voltage (mV) | 3.78 ± 0.64 | 3.87 ± 0.69 |
| Pulse width (µs) | 64.2 ± 9.96 | 62 ± 6.32 |
| Frequency (Hz) | 131 ± 6.69 | 130 ± 1.58 |
| Interleaving | **3.6% (3)** | **1.2% (1)** |
| Voltage (mV) | 3.6 ± 1.25 | 5.0 ± 0 |
| Pulse width (µs) | 60.0 ± 0 | 60 ± 0 |
| Frequency (Hz) | 125 ± 0 | 125 ± 0 |
| Double monopolar | **0%** | **1.2% (1)** |
| Voltage (mV) |  | 2.6 ± 0 |
| Pulse width (µs) |  | 60 ± 0 |
| Frequency (Hz) |  | 130 ± 0 |

Values are presented as mean ± SD and pertain to those recorded at the end of follow-up.
Patients are stratified by stimulation mode: monopolar, bipolar, interleaving, double monopolar. Some patients on Monopolar stimulation have low frequency, which explains a higher value for voltage.

### Mortality

Seventeen patients died during the study, accounting for a mortality rate at the end of follow-up of 16%. On average, patients died 62.1 ± 21.3 months after surgery (**Fig. 1**), at a mean age of 69.9 ± 6.0 years. The presence of disability milestones in the follow-up did not significantly alter the mortality rate (**Fig. 1**).

**Figure 1.**
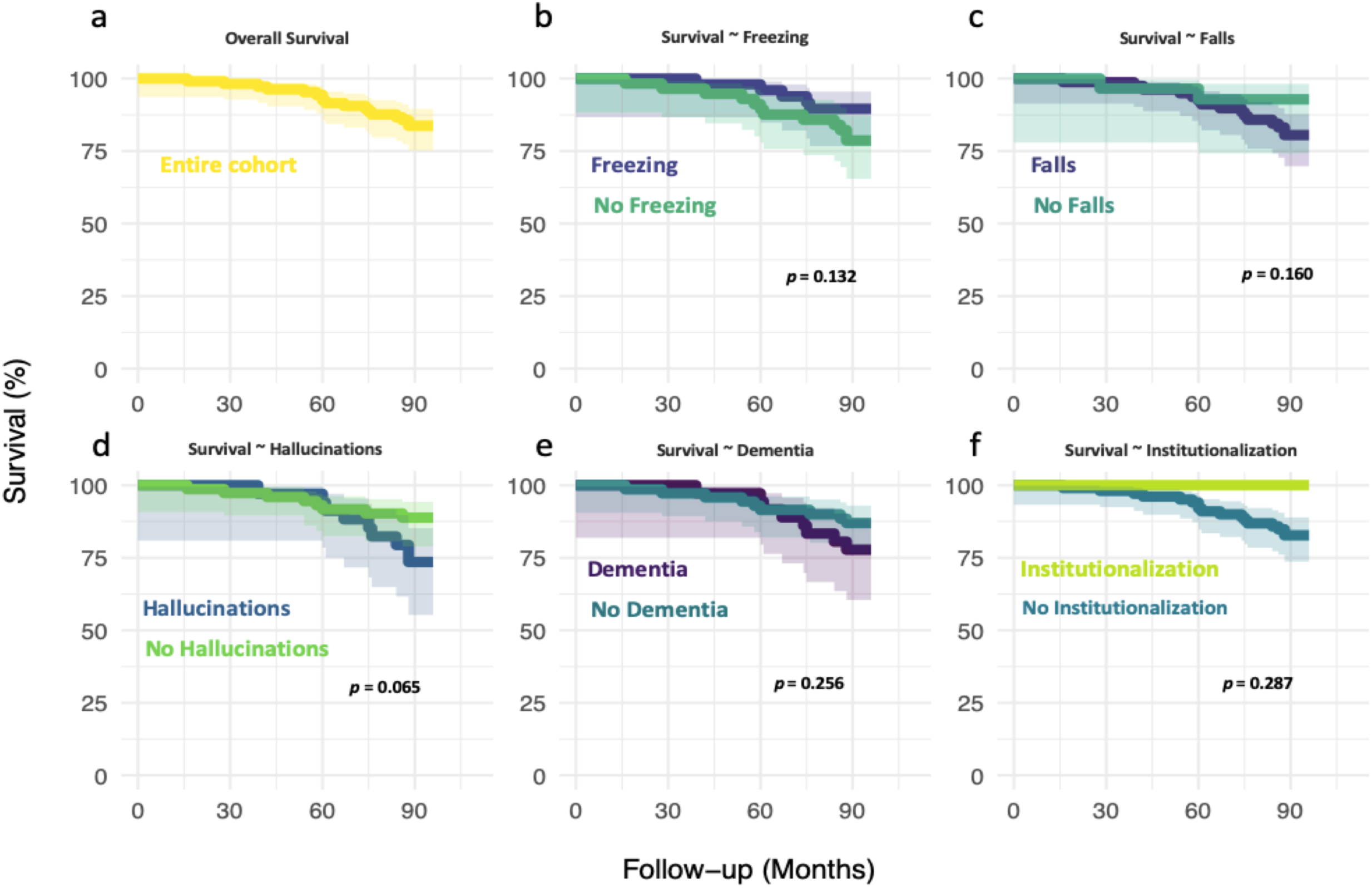
Survival plots for the entire cohort (a) and for the same cohort split between patients developing a given milestone and those failing to do so (b-f). Comparisons performed using Log-rank (Mantel-Cox) tests.

In the patients who died during follow-up, falls preceded death by 34.6 ± 13.7 months, FOG by 35.8 ± 21.4, dementia and hallucinations by 18.5 ± 14.5 and 18.7 ± 14.4, respectively. None of the patients who died were institutionalized.

Non-adjusted, univariate analysis showed that a preoperative higher UPDRS part II score in OFF (HR 1.012, C.I. 1.005-1.020, *p* = 0.001) and older age at surgery (HR 1.108, C.I. 1.008-1.218, *p* = 0.034) significantly increased mortality rates (**Sup. Fig. S2**). On multivariate analysis, age at surgery (HR 1.121, C.I. 1.035-1.463, *p* = 0.019) and the preoperative UPDRS part III score in OFF (HR 0.942, C.I. 0.890 - 0.996, *p* = 0.037) were significant drivers of overall mortality (**Fig. 3**).

### Disability Milestones

Falls were the most likely event during follow-up (**Fig. 2**), with a competing risk-adjusted probability of 73% at 8 years post-DBS (95% C.I., 0.636 - 0.897). The average time from surgery to first fall was 40.4 ± 25.4 months. FOG was observed in 47% (95% C.I., 0.377 - 0.566) of the cohort (**Fig. 2**). The average time from surgery to FOG was 39.6 ± 28.4 months. In turn, the competing risk-adjusted probability of developing hallucinations was 32% (95% C.I., 0.409 - 0.231) (**Fig. 2**), with an average time from surgery to hallucinations of 60.0 ± 20.7 months. 34% of patients developed dementia (95%C.I., 0.254 - 0.436) (**Fig. 2**), with an average time from surgery to dementia diagnosis of 56.2 ± 21.2 months. Only 6% of the cases were institutionalized (95% C.I., 0.024 - 0.117) (**Fig. 2**), after 62.3 ± 22.0 months of surgery.

**Figure 2.**
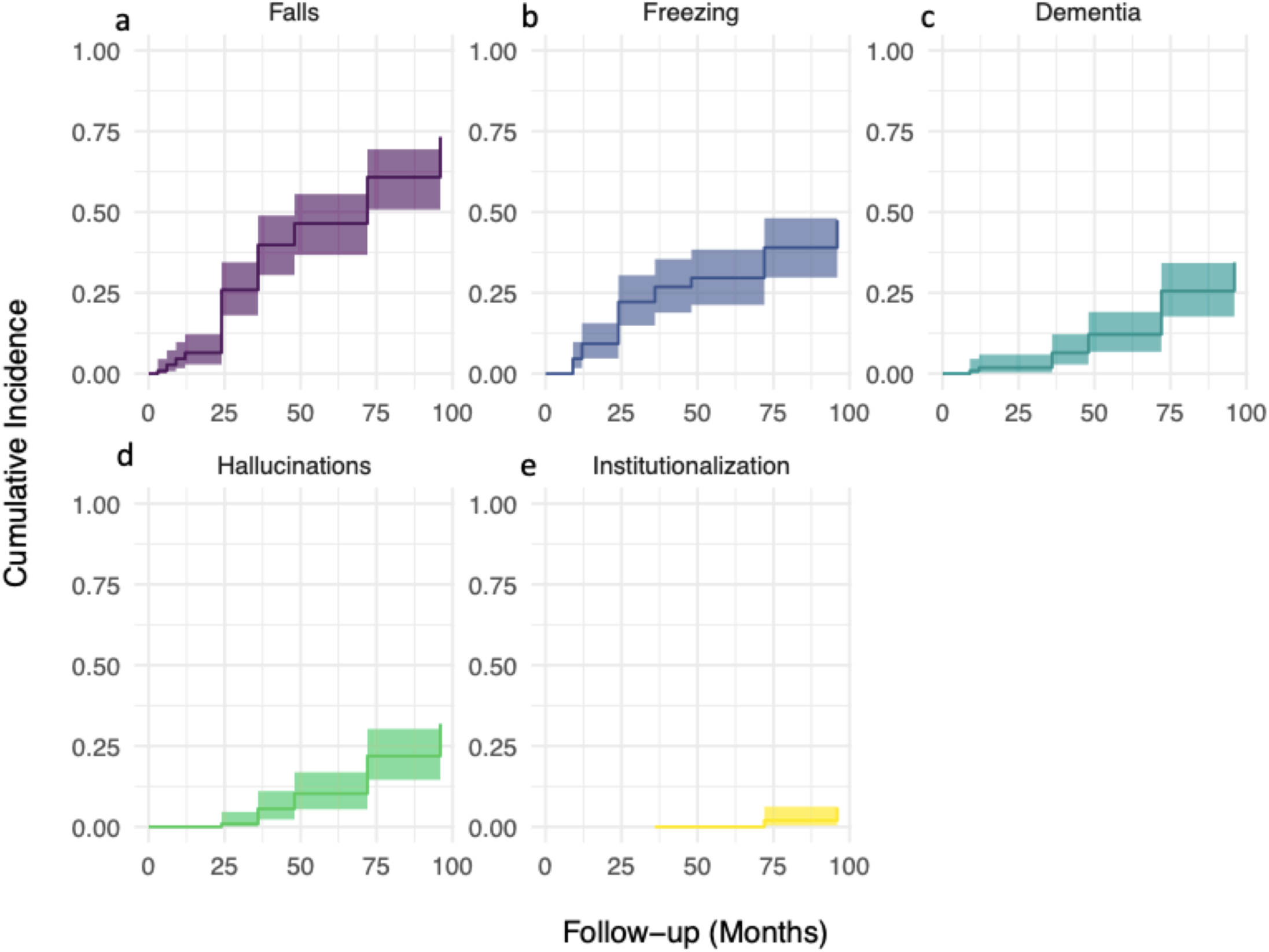
Estimated cumulative incidence curves (a-e) for each milestone event with mortality treated as competing risk. Shaded areas represent (upper and lower) 95% confidence intervals.

### Determinants of axial milestones

Fallers had worse baseline scores in UPDRS II (28.0 ± 38.3 vs 19.9 ± 6.7, p=0.015) and UPDRS III item 30 (postural instability) (1.6 ± 1.0 vs 1.0 ± 1.1, p= 0.022) in OFF than non-fallers **(Sup. Table S1)**. The baseline variables that in univariate analysis were significantly associated with falls are show in **Sup. Fig. S2.** When adjusting for confounders, older age at surgery (HR 1.184, 95% C.I., 0.779 - 1.086, *p* = 0.004) remained significant associated with falls (**Fig. 3**). The cumulative incidence of falls was significantly higher among patients who also developed FOG during follow-up (86%, 95% C.I., 0.718 – 0.932) compared to those who did not (62%, 95% C.I., 0.478 – 0.735, *p* = 0.010) and in those who developed hallucinations (85% %, 95% C.I., 0.67-0.94 vs 67%, 95% C.I., 0.55-0.77, *p* = 0.039) (**Fig. 4**).

**Figure 3.**
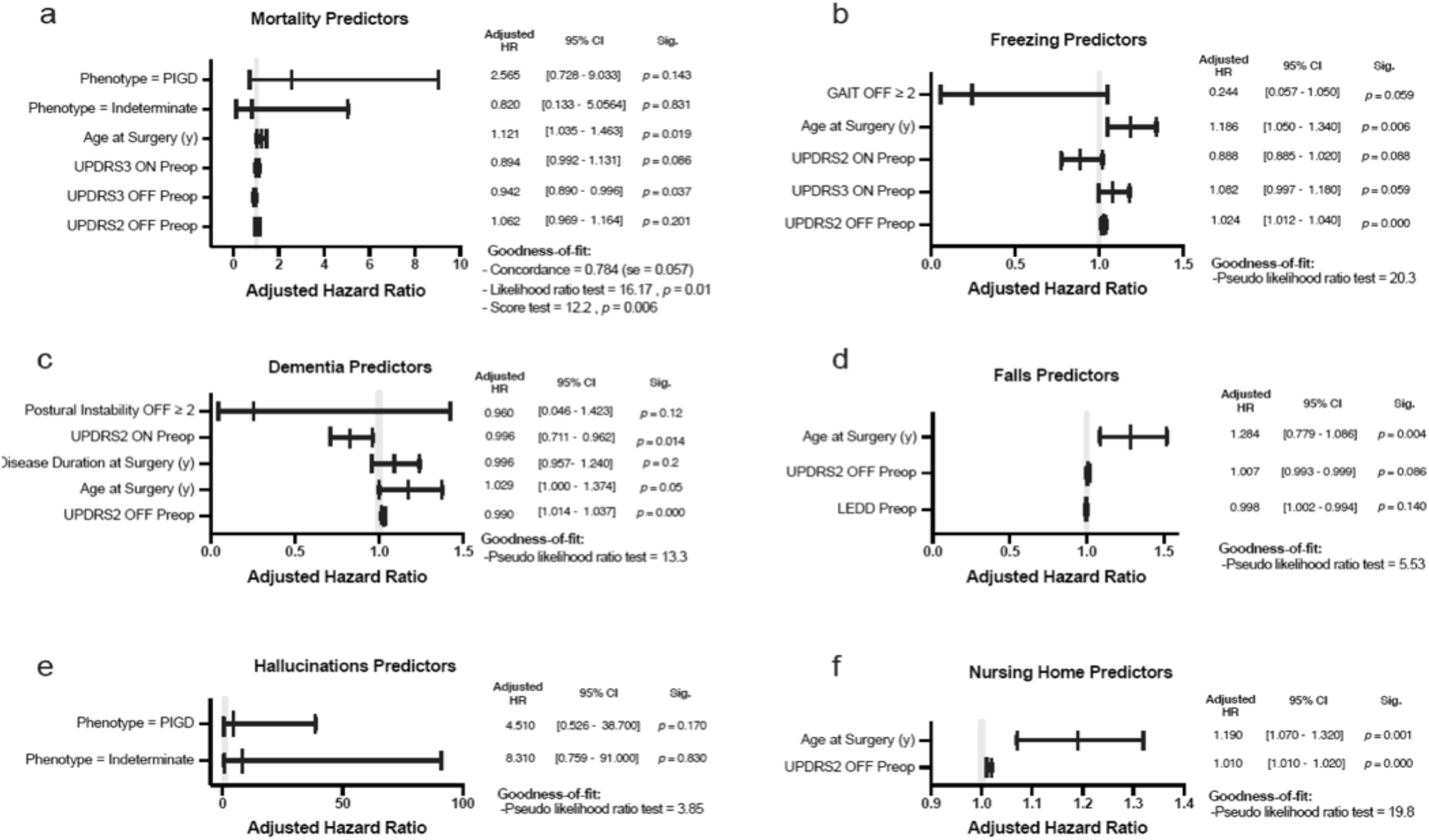
Forest plots depicting the Hazard Ratios from a Cox proportional-Hazards regression model (a, mortality) and several sub-distribution hazard models (b-f, milestone events). Each and every single variable is adjusted for all other variables in the respective plot. Multivariable regression analysis in the presence of competing risks was performed using the (semiparametric proportional hazards model) sub-distribution hazards of cumulative incidence functions based on the Gray’s test. A backward elimination method for selection of variables was using a cut-off of 0.2.

**Figure 4.**
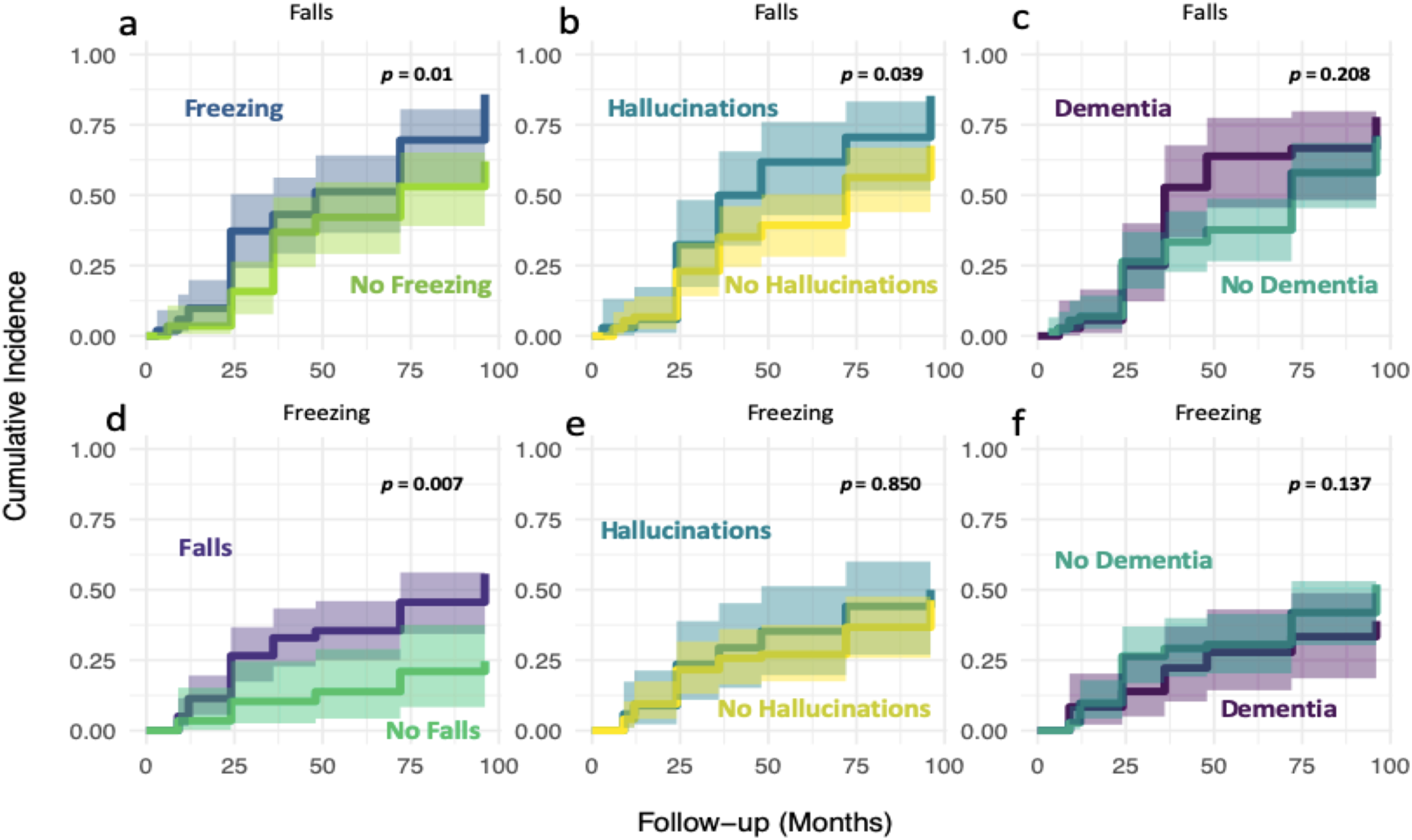
Estimated cumulative incidence plots (a-f) of falls and freezing, with equality between plots based on the Gray’s test (i.e. comparison of the weighted averages for the sub-distribution hazards of each event of interest)

Freezers had significantly higher baseline scores in UPDRS III item 29 (gait) in OFF than non-freezers (2.0 ± 0.9 vs 1.6 ± 1.5, *p* = 0.029) **(Sup. Table S2).** Univariate analysis showed that longer disease duration at surgery (HR 1.070, 95% C.I., 1.010 - 1.140, *p* = 0.021), higher preoperative UPDRS part III score in ON (HR 1.080, 95% C.I. 1.020 - 1.140, *p* = 0.012) and higher preoperative UPDRS part II score in OFF (HR 1.010, 95% C.I. 1.010 - 1.020, *p* = 0.000) significantly modulated the rate FOG **(Sup. Fig. S2).** When adjusting for confounders, older age at surgery (HR 1.186, 95% C.I. 1.050 – 1.340, *p* = 0.006) and higher UPDRS part II score in OFF (HR 1.024, 95% C.I., 1.012 - 1.040, *p* = 0.000) were significantly associated with freezing. The cumulative incidence of freezing was significantly higher in the fallers (56%, 95% C.I., 0.440 – 0.659) when compared to the non-fallers (25%, 95% C.I., 0.106 – 0.416, *p* = 0.007) (**Fig. 4**).

### Determinants of hallucinations, dementia and institutionalization

The preoperative UPDRS part II score in OFF (HR 1.010, C.I. 1.010 - 1.020, *p* = 0.000) significantly modulate the rate of hallucinations, dementia and institutionalization in univariate, non-adjusted analysis (**Sup. Fig. S2**). After adjustment for confounders, no variable remained significantly associated with hallucinations. As for dementia, both older age at surgery (HR 1.029, C.I. 1.000 - 1.374, *p* = 0.05) and preoperatory UPDRS part II score in OFF (HR 1.026, C.I. 1.014 - 1.037, *p* = 0.000) and ON (HR 0.996, C.I. 0.711 - 0.962, *p* = 0.014) significantly modulated the rate of event development. In the case of institutionalization, multivariate analysis showed that both preoperatory higher UPDRS part II score in OFF (HR 1.010, 95% C.I., 1.010 - 1.020, *p* = 0.000) and older age at surgery (HR 1.190, C.I. 1.070 - 1.320, *p* = 0.001) were significant drivers of institutionalisation(**Fig. 3**).

## Discussion

The mortality and incidence of major disability milestones of 109 PD patients followed-up for 8 years after STN-DBS have been studied using a competitive risk analysis. This cohort had an excellent benefit from DBS, with a 50% reduction in the LEDD at the end of follow-up. Mortality rate was low (overall 16%), with survivors presenting a mean age of 69 and mean disease duration of 22. For those who died, the mean time to death was about 5 years after surgery. Falls developed in three-quarters of patients, whereas FOG afflicted nearly half. Mean time to falls and FOG after surgery was about 3 years. About one-third of the patients developed hallucinations and dementia, on average 5 years after STN-DBS. Nursing home institutionalization occur in a small percentage of patients and took place on average 5 years after DBS

### Mortality

Across different studies, highly variable mortality rates have been reported (from 17 to 61%) in long-term DBS cohorts, ^10, 13, 33–36 13^ probably reflecting high heterogeneity of study populations regarding disease and follow-up duration. We have herein observed figures for cumulative mortality and cumulative incidence of falls, dementia, and hallucinations similar to those recently reported by Mahlknecht *et al.*^36^ for STN-DBS, who found a mortality rate of 17% after 7 years of follow-up. Age and disease duration at surgery were similar in both studies.^36^ Likewise, a similar survival was found after 7 years of follow-up in a cohort with a similar age at surgery.^35^ The importance of comparing cohorts with similar demographic characteristics is reinforced by the increase in mortality rates with older age at surgery.^35, 37^ Older age at PD onset has been associated with increased mortality,^38, 39^ however, findings from Kempster *et al.* have instead suggested that death (and disability milestones) occurs at around the same age regardless of the age at PD onset.^19, 40^ In our cohort, the mean age of death is around 70 years old, similar to what has been previously observed despite the longer disease duration.^19, 40^ Thus, mortality would be related to biological age more than to disease duration, or age of disease-onset.

Interestingly, regarding the milestone-to-death interval, we have found a similar temporal relationship as previous studies,^19^ reinforcing the idea that the late phase of PD (independently of age-of-onset, disease duration and DBS) follow a stereotyped pattern, with the appearance of milestones preceding death.

### Falls and FOG

Falls and FOG were both the earliest and the most frequent disability milestones and tended to occur at around the same time after surgery. The incidence of falls surpassed that of freezing in the long run. In previous studies, falls were present in 61% of a DBS cohort followed-up during > 8 years^36^ and in 64% of patients 7 years after surgery.^37^ In the same study, 64% of the patients also developed FOG.^37^ Different sample sizes and methods used to assess FOG and falls could explain the different rates between studies. A time-depended deterioration of axials signs after surgery^10, 11, 14, 16, 37, 41^ has been suggested and can explain the lower incidence of falls (32% of 260 patients) in a study with shorter disease duration at surgery (6 ± 3.8 years) and shorter follow-up time (median 3.1 years). ^42^

Older age at STN-DBS was a strong driver of increase rates of both falls and freezing. Several studies have shown an association between age and the occurrence of falls and freezing^33, 34, 42, 43^. Besides age at DBS, only a worse baseline UPDRS II OFF score was an independent predictor of freezing, but not falls. Our results reinforce the previously highlighted importance of baseline disease severity, namely in OFF, for future incidence of axial disability milestones, irrespectively of levodopa responsiveness.^6,11^ Interesting, though, is the finding that the impairment on ADL in the OFF state is a predictor of axial disability milestones. When selecting patients for DBS, special attention should be given to the performance of patients in ADL in the OFF state, besides their objective motor disability, since it can help to identify patients with a higher risk of developing disability milestones.

Even though FOG is not often assessed when studying disability milestones in advanced disease,^17, 19, 40^ we have decided to evaluate freezing because it is a symptom associated with significant disability and deterioration in QoL, and most often unresponsive to levodopa.^44^ The response of FOG to DBS-STN has been controversial^45^ and as such, there is a need for future studies to accurately characterize its response to DBS.

### Hallucinations and Dementia

Cognitive/behavioral milestones had a lower incidence than axial symptoms in our study. The rates of hallucinations and dementia after STN-DBS have been highly variable across studies, ranging from 18 to 61% for hallucinations,^15, 33, 34, 36, 37, 46^ and from 5 to 61% for dementia.^13, 16, 33, 36, 46^ An association between hallucinations and dementia (i.e., similar incidence and close temporal relationship) has been previously reported,^17, 19^ pointing to a common pathophysiology. On the one hand, a higher cortical burden of Lewy body pathology as well as a higher load of Alzheimer’s disease neuropathological changes have been associated with both dementia and hallucinations.^19, 40^ On the other hand, the presence of cortical Lewy bodies appears not to be correlated with motor milestones, indicating that cortical pathology is specifically implicated in the development of cognitive milestones.^46^

Likewise, older age is a risk factor for dementia in PD,^33, 34, 42, 43^ which might explain the higher risk of dementia in those patients older at STN-DBS.

### Nursing home placement

We found a much lower rate of admission to nursing home facilities compared to other post-DBS^16, 34, 36, 37^ and non-DBS studies^47, 48^ Nonetheless, a previous study of Portuguese late-stage non-DBS patients also found a lower frequency of nursing home admissions than that reported in the Swedish and Dutch counterparts.^48^ Sociocultural stigma together with inadequate social support systems are likely to explain such lower institutionalization rates.^36, 49^

The present study lacks a control group, but comparing the observed numbers with comparable size cohorts,^21, 50^ one is lead to conclude that STN-DBS patients developed the same disability milestones than non-DBS PD patients, but at a lower rate despite similar disease duration.^21, 49, 50^ One possible explanation is the difference in age at PD onset, with DBS patients being relatively younger at disease onset (47 vs 54, 56, and 61 years old at disease onset) and at the end of follow-up.^21, 49, 50^ Accordingly, a negative correlation between age at disease onset and time-to-development of the first disease milestone has been previously reported.^17, 19^ When compared to the Sydney cohort, a delay of 10 to 15 years in achieving the same level of disability milestones was found in the Toronto study.^37^ A younger age of onset in the Toronto study was the explanation for the difference in findings, suggesting that the disease progresses relatively slowly in younger patients but disability milestones are reached at similar ages independently of age at disease onset.^19, 37, 40^ Our data supports this hypothesis.

Interestingly, even though STN-DBS patients are a particular sample of PD patients, considering that more frequently includes patients with younger age at onset, highly responsive to levodopa and no dopaminergic resistant axial symptoms before surgery, they still end up developing the same disability milestones (**Fig. 5**). Additionally, MC have been well-controlled with STN-DBS, this study shows that surgery does not prevent the development of such milestones.

**Figure 5.**
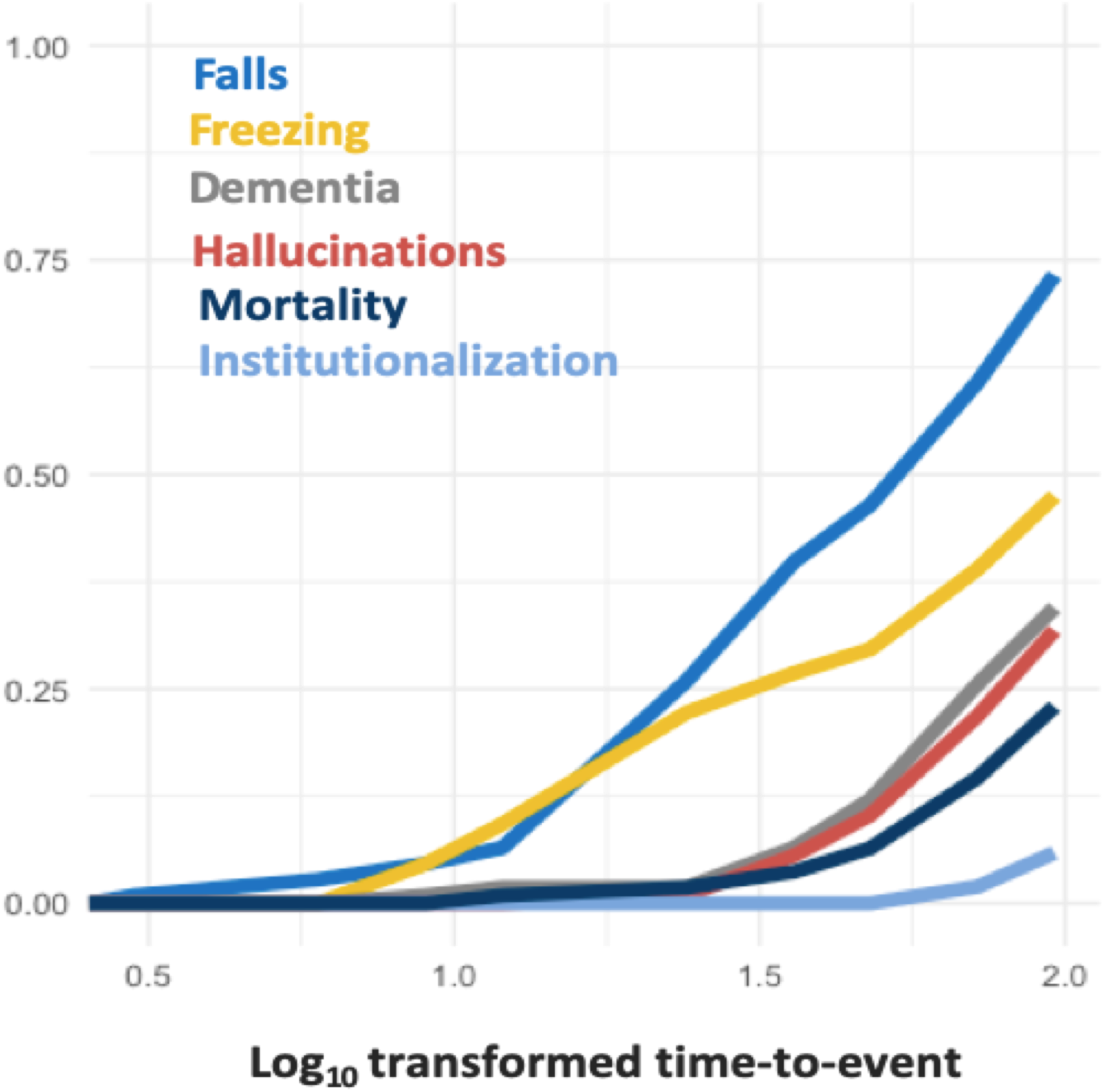
Summary incidence plot across all events under study adjusted for competing risks (falls, freezing, hallucinations, dementia, mortality, and institutionalization). For the sake of visibility, values on the X-axis correspond to the base 10 logarithmic of the number of elapsed months to event (1 -> 10 months, 1.5 -> 32 months, 2 -> 100 months).

In contrast to the usual sequence of events reported in late-stage non-DBS PD patients,^17, 19^ we have observed the development of falls antedating that of hallucinations. The decrease in dopaminergic medication after surgery and the lack of cognitive impairment before DBS may explain such a difference.^36^

## Strength and limitations

The present study is, to the best of our knowledge, the largest long-term cohort of STN-DBS PD patients comprising incidence rates and predictors of PD disability milestones. The relatively comprehensive baseline characterization has allowed for the dissection of pre-surgical clinical and demographic variables of relevance for the development of disability milestones after STN-DBS surgery. However, the retrospective nature of the study precludes a systematic evaluation of the patients regarding assessment time and instruments used. To minimize errors inherent to this type of design, we chose very clear and objective definitions for the identification of disability milestones. Like most long-term DBS follow-up studies, our study lacks a control group. To minimize this limitation, we tried to discuss our finding using historical DBS and non-DBS cohorts previously reported in the literature.

## Conclusions

Long-term mortality rate is low after STN-DBS, with older age at surgery and baseline motor impairment being the best predictors thereof. Disability milestones follow a consistent progression (mostly at very late disease stages) and tend to cluster together right before death, suggesting the same neurodegenerative process dictating disease progression in non-DBS patients to also be the predominant one among DBS patients.^15, 17, 19^ With biological age as the most important driver, there is a case to be made in support of STN-DBS surgery being performed earlier rather than later.

## Authors contribution

RB: Conceptualization, Data curation, Formal Analysis, Investigation, Methodology, Writing-original draft preparation, Writing – Review & Editing; LCG: Data curation, Writing – Review & Editing; MBC: Data curation, Writing – Review & Editing; PPL: Data curation, Writing – Review & Editing; ACC: Data curation, Writing – Review & Editing; MF: Conceptualization, Methodology, Data curation, Writing – Review & Editing; PB: Data curation, Formal Analysis, Investigation, Methodology, Software, Writing-original draft preparation, Writing – Review & Editing; AV: Data curation, Writing – Review & Editing; HC: Data curation, Writing – Review & Editing; LA: Data curation, Writing – Review & Editing; SR: Data curation, Writing – Review & Editing; A GF: Data curation, Writing – Review & Editing; JJF: Data curation, Writing – Review & Editing; MMR: Data curation, Writing – Review & Editing; MC: Conceptualization, Data curation, Formal Analysis, Investigation, Methodology, Writing-original draft preparation, Writing – Review & Editing

## Ethics approval

The study was approved by the Local Ethical Committee (“ Comissão de Ética do Centro Académico de Medicina de Lisboa – CAML”).

**Supplementary Figure S1.** Graphical Spearman correlation matrix of clinical variables of interest with r score (**a**) and p-values (**b**).

**Supplementary Figure S2 –** Unadjusted univariate analysis of predictors for mortality (A) and disability milestones (B-F). Forest plots depicting the Hazard Ratios from a Cox proportional-Hazards regression model.

## Data Availability

All data produced in the present study are available upon reasonable request to the authors

---

**Supplementary Table S1.** Demographic and clinic characteristics of fallers vs non-fallers.

|  |  |  |  |
| --- | --- | --- | --- |
|  | Fallers (n = 79) | Non-Fallers (n=30) | p-value |
| Gender: male, n=109 | 39 | 20 | 0.105 |
| Age at surgery ($\bar{x} \pm SD$), n=109 | 61.4 ± 7.0 | 60.8 ± 8.7 | 0.905 |
| Disease duration at surgery ($y \pm SD$), n=104 | 13.9 ± 5.4 | 13.5 ± 5.9 | 0.694 |
| UPDRS I ($\bar{x} \pm SD$), n=98 | 2.6 ± 1.5 | 2.6 ± 1.9 | 0.564 |
| UPDRS II OFF-MED ($\bar{x} \pm SD$), n=98 | 28.0 ± 38.3 | 19.9 ± 6.7 | 0.015 |
| UPDRS II ON-MED ($\bar{x} \pm SD$), n=98 | 9.3 ± 6.5 | 8.3 ± 4.1 | 0.928 |
| UPDRS III OFF-MED ($\bar{x} \pm SD$), n=105 | 44.7 ± 13.5 | 43.9 ± 13.3 | 0.886 |
| UPDRS III ON-MED ($\bar{x} \pm SD$), n=105 | 19.1 ± 7.8 | 18.2 ± 7.2 | 1.00 |
| H&Y OFF MED ($\bar{x} \pm SD$), n=105 | 2.8 ± 1.1 | 2.6 ± 0.9 | 0.182 |
| H&Y ON MED ($\bar{x} \pm SD$), n=105 | 2.0 ± 0.2 | 1.9 ± 0.3 | 0.066 |
| Levodopa % response ($\bar{x} \pm SD$)n=106 | 57.5 ± 13.2 | 58.0 ± 14.5 | 0.686 |
| LEDD mg ($\bar{x} \pm SD$) , n=107 | 1238.8 ± 507.7 | 1288.3 ± 561.6 | 0.687 |
| Item 29 UPDRS III OFF MED ($\bar{x} \pm SD$) , n=105 | 1.9 ± 1.6 | 1.6 ± 1.2 | 0.473 |
| Item 30 UPDRS III OFF MED ($\bar{x} \pm SD$), n=105 | 1.6 ± 1.0 | 1.0 ± 1.1 | 0.022 |
| Item 29 UPDRS III ON MED ($\bar{x} \pm SD$), n=105 | 0.4 ± 0.6 | 0.2 ± 0.5 | 0.112 |
| Item 30 UPDRS III ON MED ($\bar{x} \pm SD$), n=105 | 0.5 ± 0.6 | 0.3 ± 0.5 | 0.384 |
| Phenotype, n=95 |  |  |  |
| Tremor | 30 | 15 | 0.442 |
| PIGD | 30 | 9 |  |
| Indeterminate | 9 | 2 |  |
| MMSE score ($\bar{x} \pm SD$), n=100 | 27.9 ± 1.8 | 27.7 ± 2.7 | 0.763 |
| Neuropsychological diagnosis, n=90 |  |  |  |
| Normal | 51 | 20 | 0.272 |
| Mild Cognitive impairment | 16 | 3 |  |

Values are presented as mean ± SD. LEDD: levodopa equivalent daily dose; PIGD: postural instability gait disorder; MMSE: mini mental state examination; UPDRS: Unified Parkinson Disease Rating Scale; MDS-UPDRS: MDS-Unified Parkinson's Disease Rating Scale

**Supplementary Table S2.** Clinical and demographic characteristics of *freezers* vs non-*freezers*.

|  |  |  |  |
| --- | --- | --- | --- |
|  | FOG Patients (n = 51) | No-FOG patients (n=58) | p-value |
| Gender: male, n=109 | 30 (58.8%) | 29 (50%) | 0.356 |
| Age at surgery ($\bar{x} \pm SD$), n=109 | 62.2 ± 6.6 | 60.4 ± 8.1 | 0.224 |
| Disease duration at surgery ($y \pm SD$), n=104 | 13.88 ± 4.1 | 13.7 ± 6.7 | 0.358 |
| UPDRS I ($\bar{x} \pm SD$), n=98 | 2.7 ± 1.6 | 2.4 ± 1.7 | 0.115 |
| UPDRS II OFF-MED ($\bar{x} \pm SD$), n=98 | 23.0 ± 6.3 | 28.3 ± 45.4 | 0.324 |
| UPDRS II ON-MED ($\bar{x} \pm SD$), n=98 | 8.2 ± 6.2 | 9.7 ± 5.7 | 0.127 |
| UPDRS III OFF-MED ($\bar{x} \pm SD$), n=105 | 44.9 ± 12.2 | 44.0 ± 14.5 | 0.486 |
| UPDRS III ON-MED ($\bar{x} \pm SD$), n=105 | 17.9 ± 7.2 | 19.7 ± 7.9 | 0.217 |
| H&Y OFF MED ($\bar{x} \pm SD$), n=105 | 2.9 ± 1.0 | 2.7 ± 1.0 | 0.241 |
| H&Y ON MED ($\bar{x} \pm SD$), n=105 | 2.0 ± 0.3 | 2.0 ± 0.2 | 0.104 |
| Levodopa % response ($\bar{x} \pm SD$)n=106 | 59.6 ± 13.3 | 56.3 ± 13.3 | 0.133 |
| LEDD mg ($\bar{x} \pm SD$) , n=107 | 1320.6 ± 524.1 | 1190.8 ± 515.4 | 0.204 |
| Item 29 UPDRS III OFF MED ($\bar{x} \pm SD$) , n=105 | 2.0 ± 0.9 | 1.6 ± 1.5 | 0.029 |
| Item 30 UPDRS III OFF MED ($\bar{x} \pm SD$), n=105 | 1.3 ± 1.0 | 1.5 ± 1.1 | 0.268 |
| Item 29 UPDRS III ON MED ($\bar{x} \pm SD$), n=105 | 0.3 ± 0.6 | 0.3 ± 0.6 | 0.414 |
| Item 30 UPDRS III ON MED ($\bar{x} \pm SD$), n=105 | 0.5 ± 0.7 | 0.4 ± 0.5 | 0.624 |
| Phenotype, n=95 |  |  | 0.680 |
| Tremor | 22 | 23 |  |
| PIGD | 20 | 19 |  |
| Indeterminate | 4 | 7 |  |
| MMSE score ($\bar{x} \pm SD$), n=100 | 27.9 ± 1.5 | 27.7 ± 2.5 | 0.610 |
| Neuropsychological diagnosis, n=90 |  |  | 0.334 |
| Normal | 35 | 36 |  |
| Mild Cognitive impairment | 7 | 12 |  |

Values are presented as mean ± SD. LEDD: levodopa equivalent daily dose; PIGD: postural instability gait disorder; MMSE: mini mental state examination; UPDRS: Unified Parkinson Disease Rating Scale; MDS-UPDRS: MDS-Unified Parkinson's Disease Rating Scale

