## Supplementary figures and images for "Long-term follow-up of subthalamic nucleus deep brain stimulation in patients with Parkinson’s disease: an analysis of survival and disability milestones"

### Supp Fig 1

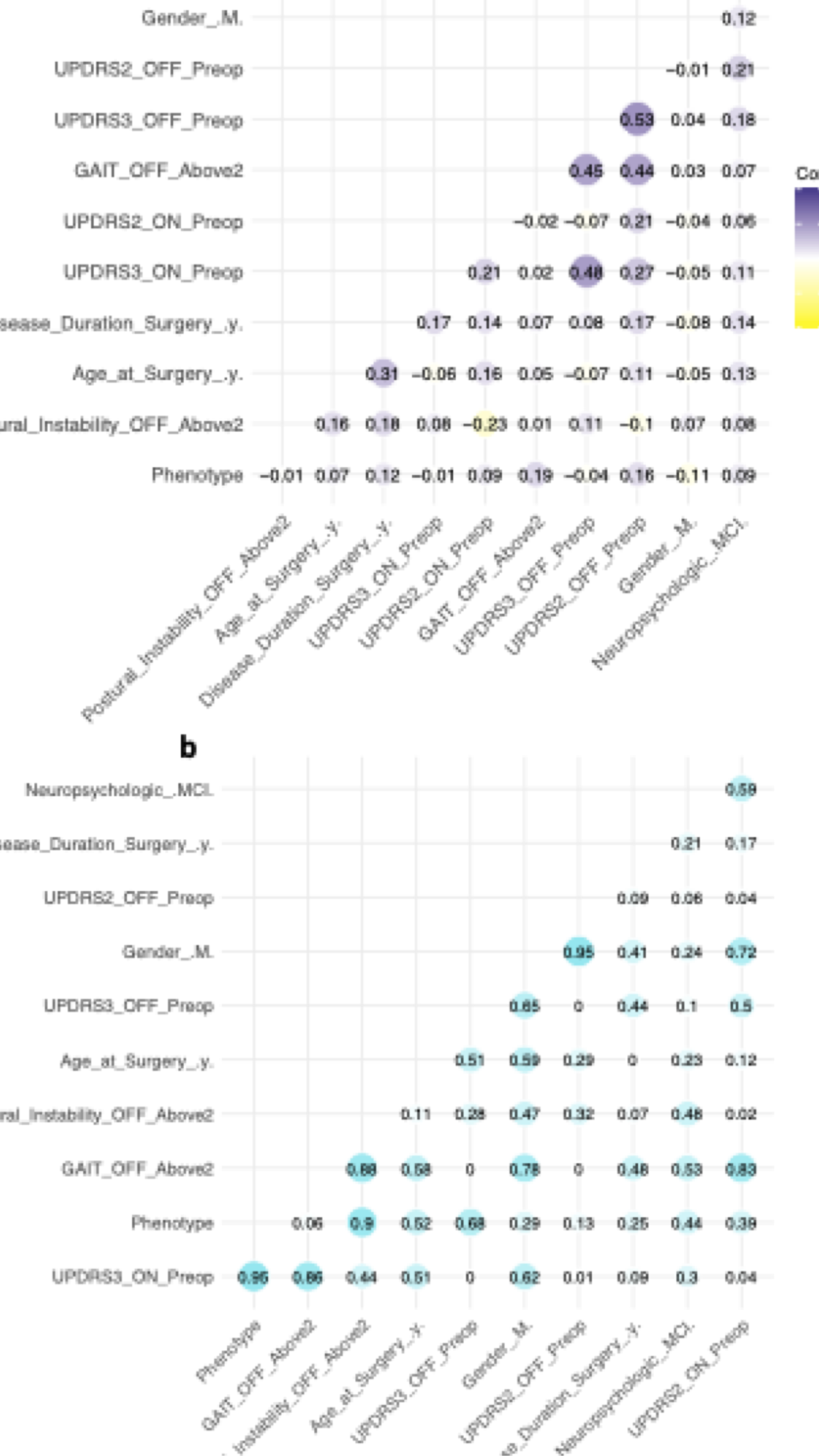

### Supp Fig 2

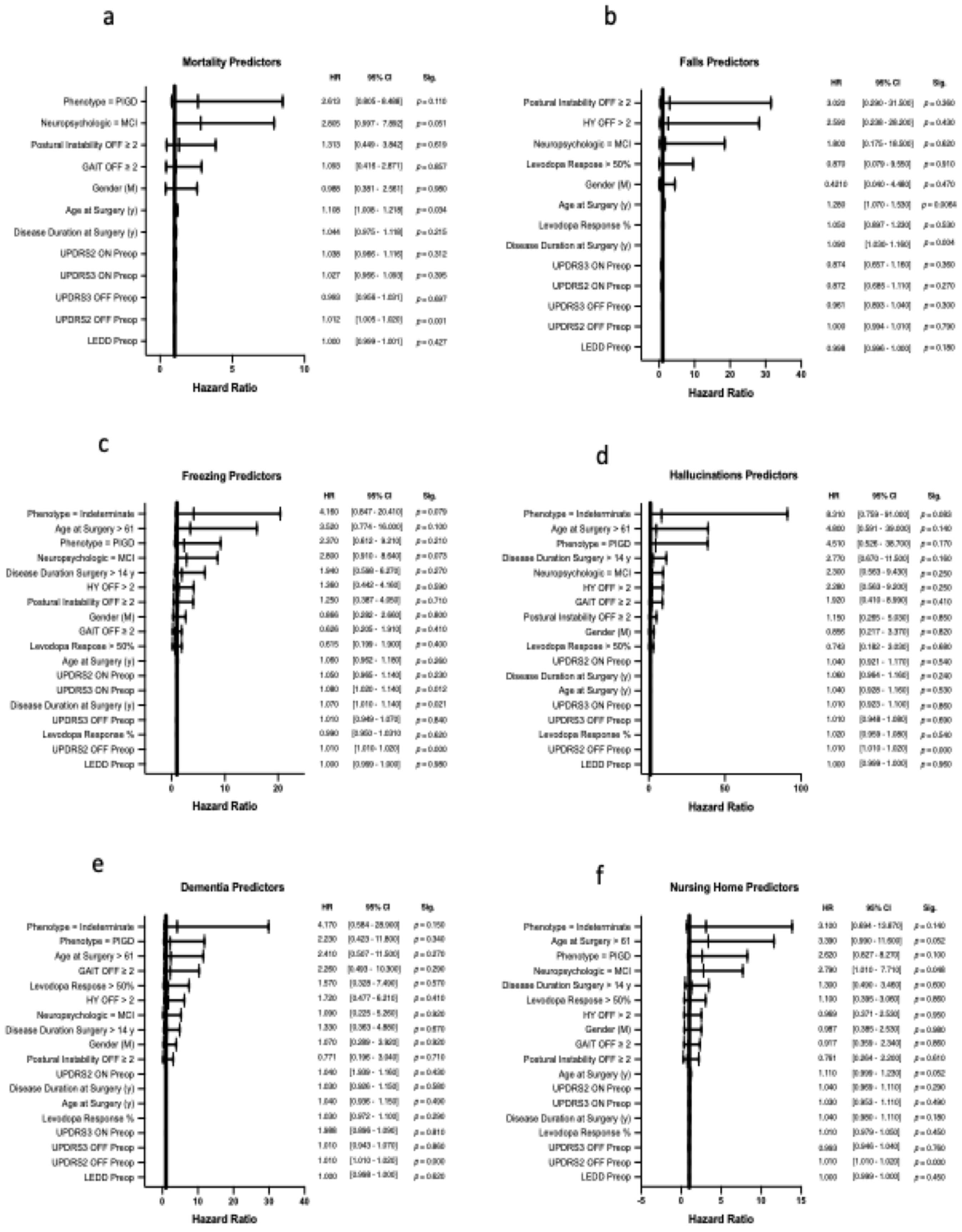
