## Supplementary material for "Long-term follow-up of subthalamic nucleus deep brain stimulation in patients with Parkinson’s disease: an analysis of survival and disability milestones": Supp Table 1

| Supplementary Table S1 – Demographic and clinic characteristics of fallers vs non-fallers | | | |
| --- | --- | --- | --- |
|  | Fallers (n = 79) | Non-Fallers (n=30) | *p-*value |
| Gender: male, n=109 | 39 | 20 | 0.105 |
| Age at surgery (x̅ ± SD), n=109 | 61.4 ± 7.0 | 60.8 ± 8.7 | 0.905 |
| Disease duration at surgery (y ± SD), n=104 | 13.9 ± 5.4 | 13.5 ± 5.9 | 0.694 |
| UPDRS I (x̅ ± SD), n=98 | 2.6 ± 1.5 | 2.6 ± 1.9 | 0.564 |
| UPDRS II OFF-MED (x̅ ± SD), n=98 | 28.0 ± 38.3 | 19.9 ± 6.7 | 0.015 |
| UPDRS II ON-MED (x̅ ± SD), n=98 | 9.3 ± 6.5 | 8.3 ± 4.1 | 0.928 |
| UPDRS III OFF-MED (x̅ ± SD), n=105 | 44.7 ± 13.5 | 43.9 ± 13.3 | 0.886 |
| UPDRS III ON-MED (x̅ ± SD), n=105 | 19.1 ± 7.8 | 18.2 ± 7.2 | 1.00 |
| H&Y OFF MED (x̅ ± SD), n=105 | 2.8 ± 1.1 | 2.6 ± 0.9 | 0.182 |
| H&Y ON MED (x̅ ± SD), n=105 | 2.0 ± 0.2 | 1.9 ± 0.3 | 0.066 |
| Levodopa % response (x̅ ± SD)n=106 | 57.5 ± 13.2 | 58.0 ± 14.5 | 0.686 |
| LEDD mg (x̅ ± SD) , n=107 | 1238.8 ± 507.7 | 1288.3 ± 561.6 | 0.687 |
| Item 29 UPDRS III OFF MED (x̅ ± SD) , n=105 | 1.9 ± 1.6 | 1.6 ± 1.2 | 0.473 |
| Item 30 UPDRS III OFF MED (x̅ ± SD), n=105 | 1.6 ± 1.0 | 1.0 ± 1.1 | 0.022 |
| Item 29 UPDRS III ON MED (x̅ ± SD), n=105 | 0.4 ± 0.6 | 0.2 ± 0.5 | 0.112 |
| Item 30 UPDRS III ON MED (x̅ ± SD), n=105 | 0.5 ± 0.6 | 0.3 ± 0.5 | 0.384 |
| Phenotype, n=95  Tremor  PIGD  Indeterminate | 30  30  9 | 15  9  2 | 0.442 |
| MMSE score (x̅ ± SD), n=100 | 27.9 ± 1.8 | 27.7 ± 2.7 | 0.763 |
| Neuropsychological diagnosis, n=90  Normal  Mild Cognitive impairment | 51  16 | 20  3 | 0.272 |
