## Supplementary material for "Long-term follow-up of subthalamic nucleus deep brain stimulation in patients with Parkinson’s disease: an analysis of survival and disability milestones": Supp Table 2

| **Supplementary Table S2 – Clinical and demographic characteristics of *freezers* vs non-*freezers*** | | | |
| --- | --- | --- | --- |
|  | FOG Patients (n = 51) | No-FOG patients (n=58) | p-value |
| Gender: male, n=109 | 30 (58.8%) | 29 (50%) | 0.356 |
| Age at surgery (x̅ ± SD), n=109 | 62.2 ± 6.6 | 60.4 ± 8.1 | 0.224 |
| Disease duration at surgery (y ± SD), n=104 | 13.88 ± 4.1 | 13.7 ± 6.7 | 0.358 |
| UPDRS I (x̅ ± SD), n=98 | 2.7± 1.6 | 2.4 ± 1.7 | 0.115 |
| UPDRS II OFF-MED (x̅ ± SD), n=98 | 23.0 ± 6.3 | 28.3± 45.4 | 0.324 |
| UPDRS II ON-MED (x̅ ± SD), n=98 | 8.2 ± 6.2 | 9.7±5.7 | 0.127 |
| UPDRS III OFF-MED (x̅ ± SD), n=105 | 44.9 ± 12.2 | 44.0 ± 14.5 | 0.486 |
| UPDRS III ON-MED (x̅ ± SD), n=105 | 17.9 ± 7.2 | 19.7 ± 7.9 | 0.217 |
| H&Y OFF MED (x̅ ± SD), n=105 | 2.9 ± 1.0 | 2.7 ± 1.0 | 0.241 |
| H&Y ON MED (x̅ ± SD), n=105 | 2.0 ± 0.3 | 2.0 ± 0.2 | 0.104 |
| Levodopa % response (x̅ ± SD)n=106 | 59.6 ± 13.3 | 56.3 ± 13.3 | 0.133 |
| LEDD mg (x̅ ± SD) , n=107 | 1320.6±524.1 | 1190.8±515.4 | 0.204 |
| Item 29 UPDRS III OFF MED (x̅ ± SD) , n=105 | 2.0 ± 0.9 | 1.6 ± 1.5 | 0.029 |
| Item 30 UPDRS III OFF MED (x̅ ± SD), n=105 | 1.3 ± 1.0 | 1.5 ± 1.1 | 0.268 |
| Item 29 UPDRS III ON MED (x̅ ± SD), n=105 | 0.3 ± 0.6 | 0.3 ± 0.6 | 0.414 |
| Item 30 UPDRS III ON MED (x̅ ± SD), n=105 | 0.5 ± 0.7 | 0.4 ± 0.5 | 0.624 |
| Phenotype, n=95  Tremor  PIGD  Indeterminate | 22  20  4 | 23  19  7 | 0.680 |
| MMSE score (x̅ ± SD), n=100 | 27.9 ± 1.5 | 27.7 ± 2.5 | 0.610 |
| Neuropsychological diagnosis, n=90  Normal  Mild Cognitive impairment | 35  7 | 36  12 | 0.334 |
